# Estimation of Heavy Metal Contamination in Selected Marine Fish in Bangladesh and Their Health Impact

**DOI:** 10.64898/2026.02.02.26345413

**Authors:** Muhammad Anisur Rahaman, Israt Jahan, Syeda Saima Alam, Lincon Chandra Shill, Asif Mahmud Dihan, Abdullah Al Mamun

**Affiliations:** Department of Food Technology and Nutrition Science, Faculty of Science, Noakhali Science and Technology University, Noakhali-3814, Bangladesh

**Keywords:** Heavy metal, Marine Fish, Bangladesh, Health impact, Estimated daily intake, Target hazard quotient, Hazard index, Target cancer risk

## Abstract

**Background:** Sea fish traditionally serves as a protein source and plays a crucial and indispensable role in providing nutrition for the people of Bangladesh. However, frequent consumption may potentially indicate health risks through contamination with toxic heavy metals. The main purpose of this study is to evaluate the levels of heavy metal concentrations (Cr, Fe, Ni, Mn, Cu, and Pb) in selected sea fish from Chattogram and Cox’s Bazar districts in Bangladesh.

**Methods:** A wet digestion technique was employed to prepare the samples for analyzing heavy metals. Atomic Absorption Spectrophotometry (AAS) in flame and furnace technique was utilized for the estimation of heavy metal content. The health risk of human was evaluated grounded on Estimated Daily Intake (EDI), Target Hazard Quotient (THQ), Total Target Hazard Quotient (TTHQ) or Hazard Index (HI), and Target Cancer Risk (CR).

**Result:** The descending chronology of average concentrations for the selected heavy metals was as follows: Fe (32.36) > Ni (12.12) > Pb (9.70) > Cu (7.29) > Mn (5.94) > Cr (5.22). The correlations (r0.587) between Cr and Mn were found significantly positive which indicated the parameters were interconnected with each other and likely have a common origin within the study area. EDI values of four samples in the case of Cr and six values for Pb exceeded the reference doses (R_f_D) which included Bombay Duck, Ilish, Silver Pomfret, Longfin Tuna, Indian Threadfin, and Bigeye Ilisha. In six sea fish samples, the THQ for Cr and Pb crossed the allowable limit of 1. The TTHQ/HI values for seven fish species were higher than 1 ranging from Bigeye Ilisha (3.25) to Indian Mackerel (1.35). The CR values for the majority of the heavy metals fell within an acceptable range.

**Conclusions:** From a public health perspective, this study revealed that continuous consumption of heavy metals, resulting in non-oncogenic and oncogenic health implications as well.

## Background

In the nutritional perspective, fish as well as fish derived products are nutrient-dense food source having a low-calorie count characterized by high quality protein [1]. The overall well-being and physiological functioning of the human body can be achieved by having a properly balanced diet that includes enough fish [2]. The seafood intake has markedly increased over the recent several years, and it now serves as a significant source of vitamins, fatty acids, and critical minerals [3-5]. Moreover, it is better to choose fish for achieving a balanced diet since it supplies all the essential amino acids, vitamins, minerals, and fatty acids. Fish provides very affordable and easily accessible protein sources (15-20%) [6]. Fish constitutes a vital component of diet and livelihoods of more than one billion people worldwide [7].

Heavy metal is a metallic element with a high atomic weight that tends to build up in the food chain and can harm living beings in low quantities [8]. The potential impact of heavy metal on food chain is a major global concern which affects bio-magnification [9, 10]. The mechanism of bioaccumulation allows fish and other marine animals to incorporate these harmful pollutants within their bodies. The most toxic heavy metals are classified as Nickel (Ni), Cadmium (Cd), Lead (Pb), Chromium (Cr), Mercury (Hg), Copper (Cu) and Arsenic (As) [11]. These components are very hazardous and may accumulate in seafood [12]. Thus, seafood consumption constitutes a principal pathway to toxic elements for human; hence, it could endanger their physical health. Eating such contaminated fish for a long time can pose in serious health injury or death [13]. Both naturally and artificially, heavy metals may occur in aquatic environment [14]. Even at low concentrations for human consumption, several heavy metals may have the tendency to bio-accumulate and result in adverse health outcome leading to serious morbidity or even mortality [15, 16]. The severity of heavy metal contamination in the ocean and seafood is increasing along with the growth of human society and the economy, which is harmful to people’s health [17]. Marine foods including fish, prawn, crab, and mussels are delicacies and essential daily consumables. The permeable body surface of the organism absorbs trace metals directly from surrounding marine water, and the gut also absorbs them from the food and the ocean [18]. The health consequences resulted from the adverse impacts of heavy metals have been extensively documented in the scientific literature, and they can include liver and kidney damage, cardiovascular illnesses and, in severe cases, even death [19, 20]. Heavy metals can be extremely toxic, and cause cancer in the human brain, prostate gland, and other organs with both short-term and long-term exposure [21-23]. The provisional tolerable weekly intake (PTWI) has been standardized by the Expert Committee on Food Additives of the Food and Agriculture Organization (FAO) and the World Health Organization (WHO) as a health-based benchmark to safeguard the consumers from the toxicological exposure to heavy metal from intake of fish. [24-26]. Chattogram and Cox’s Bazar are coastal districts of Bangladesh where residents often ingest marine fish collected from different local markets. The quantification of heavy metals in marine fish gathered from these sites is a significant public health concern. Therefore, this study targets to acquire information about the amount of heavy metals (Mn, Cr, Fe, Ni, Cu, and Pb) in sea fish from Chattogram and Cox’s Bazar districts in Bangladesh. Moreover, the study also explored the plausible health jeopardies related to heavy element consumption in human.

## Methods and Materials

### Study location

This study was carried out within the geographical regions of Chattogram and Cox’s Bazar districts in Bangladesh. The areas were chosen, as they are coastal areas of Bangladesh where sea fish and fish products are very common in regular diet.

**Figure :**
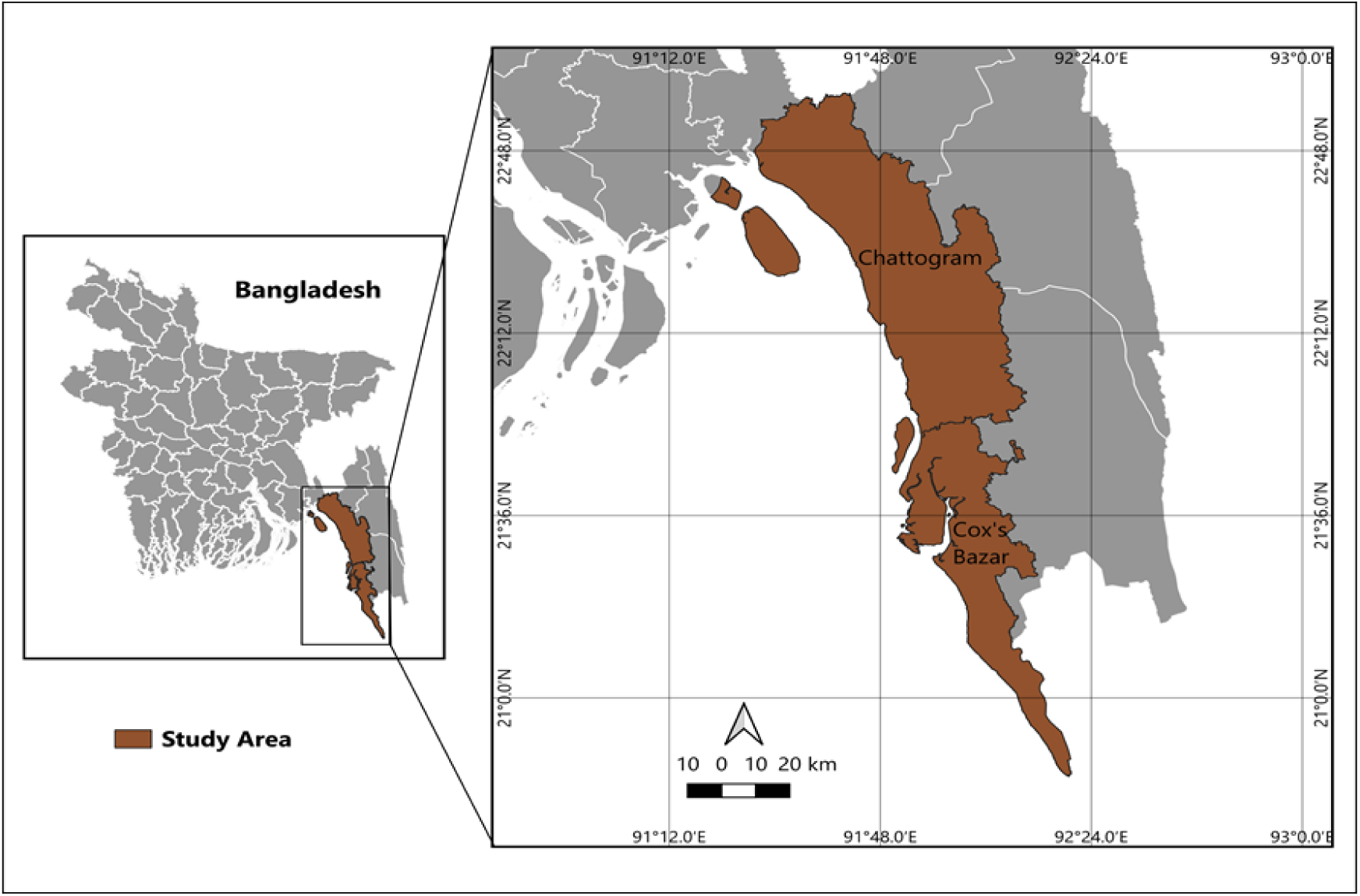
Map showing the sampling site (Chattogram and Cox’s Bazar districts in Bangladesh)

### Sample collection

Twelve commonly consumed sea fish samples were gathered from various fish marketplaces located in the Chattogram and Cox’s Bazar districts of Bangladesh. The sea fish were securely packaged in sterile polyethylene bags and promptly transferred to the research center for subsequent processing.

**Table 1:**
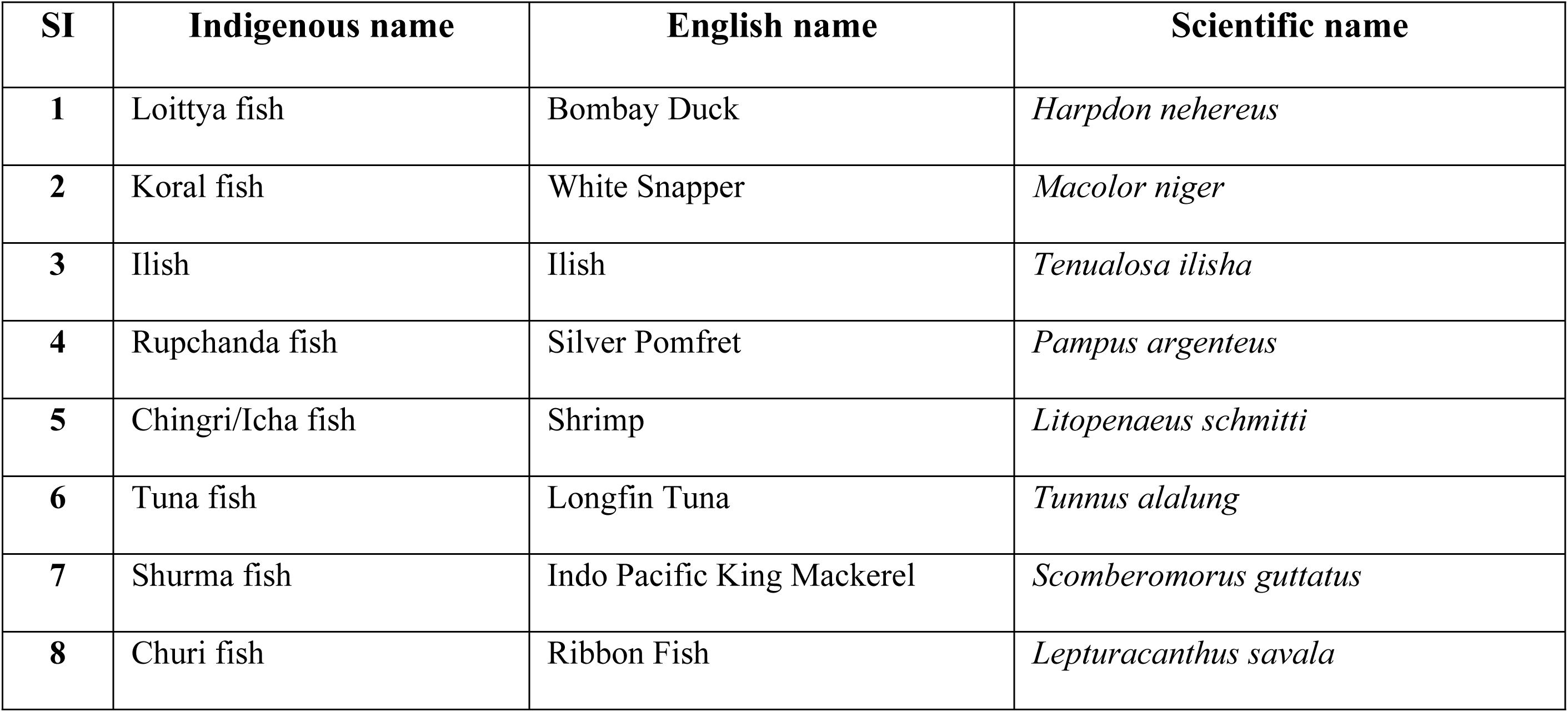
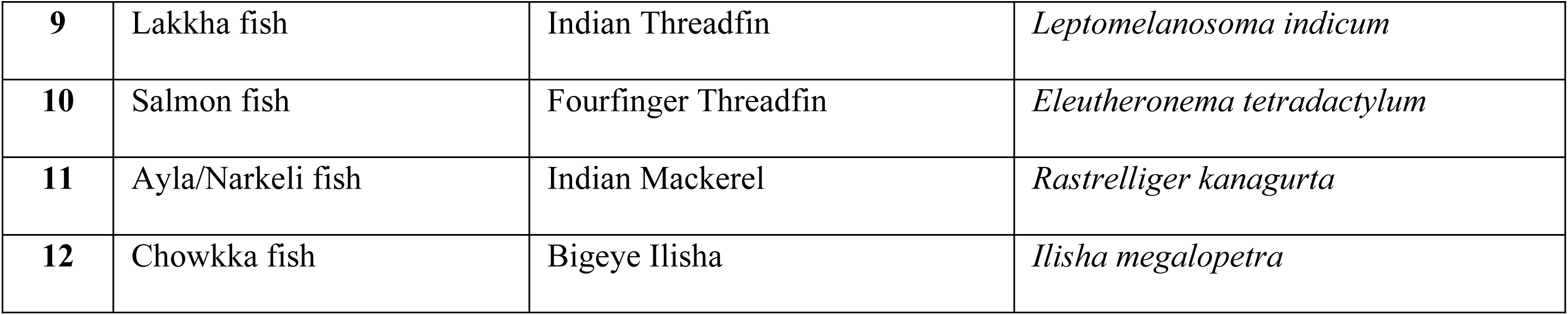
Identification of fish samples under this research.

### Wet Digestion of Sample

Cleaned and chopped samples were transferred into hot air oven at 121°C until they were crispy enough to grind. The dried sample has been transferred into a mortar to grind and homogenize the samples thoroughly for easy digestion. The homogenized samples were then kept in a desiccator for a whole night. In the wet digestion method, a mixture of HN*O*_3_ (69% concentrated) and H_2_SO_4_(95-97 % concentrated) was used in 1:4 ratio. In a volumetric flask, a homogenized, dried sample weighing 0.5g was placed. It was then completely mixed with a 5 ml acid solution. Using a heating block under thermostat control, the mixture was subsequently heated to a temperature of 170°C, where it stayed for about an hour. After that, the volumetric flask was allowed to gradually cool. After cooling down, 2 ml of 30% (w/v) H₂O₂ was added to each sample, which was then reheated unless transparent solutions were obtained. The solution was subsequently poured into a 250 ml conical flask and the sample was filtered with filter paper after adding double-distilled de-ionized water into the sample to make the sample 250ml. A sample solution volume of 50 ml was taken from the 250 ml solution and put into a falcon tube.

### Heavy Metal Analysis

The analysis for heavy metals was conducted utilizing a Perkin Elmer Inc. PinAAcle TM 900H Atomic Absorption Spectrometer (AAS), implement double-beam background correction through the utilization of high-intensity deuterium arc lamp within a continuum source, employing dual beam optics.

Employing a blank solution, the apparatus was calibrated using a "linear calibration through zero" approach. Additionally, a standard solution with defined concentrations of the particular heavy metals under this study was used. Each fish sample under analysis performed a triplicate analysis using the AAS method on cleared and digested solutions [27]. To detect each metal, specific wavelengths were employed with the use of dual-beam deuterium arc lamp for chromium, iron, nickel, manganese, copper, and lead. The calibration standards exhibited a linearity (R^2^) ranging from 0.997 to 0.999. According to a Malaysian study by Nordin et al. and Selamat et al., the limit of detection (LOD) was set at 0.001 μg/g, and the standard measuring time for an accurate determination was 3 seconds [28].

### Health Risk Assessment

The amount of heavy metals in marine fish was assessed by calculating various parameters, including the EDI, THQ, HI, and CR values of metals, separately for residents of Bangladesh.

#### EDI

The EDI is typically quantified in units of milligrams per kilogram of body weight per day (mg/kg bw/day) [29].

The mean metal content of each sample was computed, and this value was then multiplied by the corresponding consumption rate to calculate the EDI. The per day consumption rate was measured using the following equation [30].

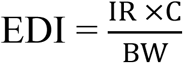

In this equation, IR represents the fish ingestion rate (kg/person/day), C signifies the heavy metal concentration found in fish samples (mg/kg), and BW denotes the average body weight (kg) of an adult individual.

#### THQ

The quantification of non-cancer risk linked to exposure to contaminants is referred to as the THQ. The THQ is computed utilizing the subsequent formula [31].

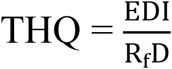

R_f_D represents the reference dose (mg/kg-bw/day) for metals determined the United States Environmental Protection Agency (USEPA). The R_f_D values for manganese (Mn), chromium (Cr), copper (Cu), nickel (Ni), and lead (Pb) are 0.14, 0.003, 0.04, 0.02, and 0.004 (mg/kg-bw/day), respectively [32-34]. In accordance with the directives outlined by the Chinese Nutrition Society (CNS), the R_f_D for iron (Fe) stands at 0.667 mg/kg-bw/day [35, 36]. When the THQ is less than 1, it suggests that individuals exposed to the contaminants are unlikely to face adverse health risks. However, if the THQ exceeds 1, it indicates a potential health concern, imposing the application of defensive and risk management measures [37].

#### HI / TTHQ

The HI was determined by aggregating the THQ values of each individual element, thereby measuring the cumulative possible health complications associated with exposure to multiple metals [38].

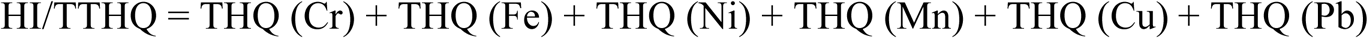

When the HI value exceeds 1, it signifies that the exposed population may be subject to an elevated risk of experiencing potential adverse health effects. Conversely, when HI is less than 1, it suggests that the population is unlikely to encounter noticeable adverse effects [39-42].

#### CR

The CR was employed to signify the risk of developing cancer. The carcinogenic risks associated with Cadmium (Cd), Chromium (Cr), Lead (Pb), and Nickel (Ni) were estimated using the formula prescribed by the United States Environmental Protection Agency (USEPA) [43].

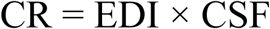

Here, CSF represents the oral carcinogenic slope factor as provided by the USEPA [43].

The provided CSF values for different heavy metals were as follows: 0.041 mg/kg/day for Cr [44], 0.6 mg/kg/day for Cd [44], 0.0017 mg/kg/day for Ni [45], and 0.0085 mg/kg/day for Pb [46].

### Statistical analysis

The statistical analyses were performed utilizing Microsoft Excel and version 25 of the statistical package for social sciences (SPSS) (p< 0.05).

## Results and discussions

### The concentrations of heavy metals in sea fish samples (mg/kg)

The mean concentration of chosen heavy metals exhibited a descending order as follows: Fe (32.36) > Ni (12.12) > Pb (9.70) > Cu (7.29) > Mn (5.94) > Cr (5.22). For each sea fish species, the findings have been presented as the mean value along with the corresponding standard deviation (SD), and these statistical summaries were provided in Table 2. There was a substantial deviation in the heavy metal concentrations across various types of sea fishes. Among them, iron contents were found in greater concentration than those of other heavy metals.

**Table 2:** The determination of heavy metals concentrations in sea fish samples (mg/kg)

| Sample name | Cr ± SD | Fe ± SD | Ni ± SD | Mn ± SD | Cu ± SD | Pb ± SD |
| --- | --- | --- | --- | --- | --- | --- |
| **1. Bombay Duck** | 1.30 ± 0.0290 | 30.00 ± 0.0034 | 12.30 ±<br>0.0007 | 2.10 ± 0.0040 | 8.00 ± 0.0109 | 12.10 ± 0.0040 |
| **2. White Snapper** | 1.70 ± 0.0109 | 38.00 ± 0.0015 | 11.80 ±<br>0.0028 | 11.00 ± 0.0017 | 5.80 ± 0.0174 | 3.90 ± 0.0146 |
| **3. Ilish** | 9.82 ± 0.0304 | 32.50 ± 0.0023 | 11.80 ±<br>0.0011 | 1.60 ± 0.0009 | 5.50 ± 0.0115 | 13.20 ± 0.0546 |
| **4. Silver Pomfret** | 9.79 ± 0.0107 | 41.70 ± 0.0401 | 12.20 ±<br>0.0009 | 17.00 ± 0.0017 | 5.20 ± 0.0400 | 12.90 ± 0.0516 |
| **5. Shrimp** | 1.91 ± 0.0298 | 34.10 ± 0.0004 | 12.40 ±<br>0.0008 | 1.20 ± 0.0009 | 11.50 ± 0.0028 | 5.20 ± 0.0262 |
| **6. Longfin Tuna** | 10.80 ± 0.0190 | 36.10 ± 0.1590 | 12.90 ±<br>0.0008 | 1.30 ± 0.0016 | 7.80 ± 0.0169 | 13.70 ± 0.0108 |
| **7. Indo Pacific King Mackerel** | 1.90 ± 0.0090 | 31.50 ± 0.0059 | 12.50 ±<br>0.0009 | 1.50 ± 0.0006 | 6.70 ± 0.0503 | 5.90 ± 0.0056 |
| **8. Ribbon Fish** | 1.95 ± 0.0205 | 39.30 ± 0.0022 | 12.70 ±<br>0.0006 | 1.10 ± 0.0006 | 5.20 ± 0.0186 | 6.20 ± 0.0134 |
| **9. Indian Threadfin** | 1.99 ± 0.3060 | 24.70 ± 0.0022 | 12.00 ±<br>0.0023 | 1.30 ± 0.0004 | 7.80 ± 0.0197 | 13.40 ± 0.0145 |
| **10. Fourfinger Threadfin** | 1.98 ± 0.0312 | 25.80 ± 0.0018 | 11.50 ±<br>0.0018 | 11.00 ± 0.0004 | 7.40 ± 0.0426 | 2.60 ± 0.0053 |
| **11. Indian Mackerel** | 1.94 ± 0.0469 | 26.80 ± 0.0027 | 11.60 ±<br>0.0017 | 1.20 ± 0.0002 | 8.30 ± 0.0245 | 14.40 ± 0.0076 |
| **12. Bigeye Ilisha** | 17.66 ± 0.0108 | 27.90 ± 0.0023 | 11.80 ±<br>0.0043 | 21.00 ± 0.0017 | 8.30 ± 0.0228 | 13.00 ± 0.016 |
| **Mean Value** | 5.22 | 32.36 | 12.12 | 5.94 | 7.29 | 9.70 |

### Chromium (Cr) in sampled sea fish (mg/kg)

The study found that the average concentration of Cr in fish samples was 5.22 mg/kg. The maximum concentration was found in Bigeye Ilisha which was 17.66 mg/kg and the lowest value was found in Bombay Duck which was 1.30 mg/kg. As per FAO guidelines, the recommended allowable range for Cr in fish falls between 0.65 to 4.35 mg/kg [47]. In this study, it is found Cr concentration of the major fish samples exceed the recommended ranges. A study led by Dhaneesh et al. found that the chromium levels in all analyzed Longfin tuna samples exceeded the established Maximum Permissible Limit (MPL) [48]. Another study found that the concentration of Cr recorded in Indo Pacific King Mackerel exceeded the MPL [49]. A recent Indian study explored that Cr concentration in Indian Mackerel exceeds the MPL [50]. Another study carried out at Chennai, India reported that the range of Cr levels was between 2.05 and 2.19 mg /kg [51].

### Iron (Fe) in sampled sea fish (mg/kg)

The mean value for Fe concentration was 32.36 mg/kg. Iron concentration in this study was the highest among all metals. The highest Fe concentration detected in Silver Pomfret fish was 41.70 mg/kg, level below the MPL(100 mg/kg) for iron in fish reported by the Joint FAO/WHO Expert Team [52]. In a comparative study, the Fe content in Silver Pomfret fish was reported 6.84 mg/kg [53]. A different study found the mean concentrations of Fe in fish in between 0.0009 to 0.001 mg/kg [51]. Also, a study in Bangladesh reported that the mean iron concentration in fish ranged from 90.37 mg/kg to 104.55 mg/kg, which was greater than the levels observed in this particular study [54].

### Nickel (Ni) in sampled sea fish (mg/kg)

The mean concentration of Ni was found 12.12 mg/kg in our current study. The Longfin Tuna has shown the highest recorded concentration at 12.9 mg/kg, while the Fourfinger Threadfin fish displayed the lowest recorded value at 11.5 mg/kg. However, these values fall within the acceptable limits established by the Joint FAO/WHO Expert report [25]. An Indian study found that the concentration of Ni ranged from 60.4 mg/kg to 69.9 mg/kg, surpassing the levels observed in the current study [51]. In a Bangladeshi study conducted by Jothi et al. the concentration of Ni in Bombay Duck extended from 1.902 mg/kg to 0.110 mg/kg, which was notably below the established permissible limit [55].

### Manganese (Mn) in sampled sea fish (mg/kg)

We have found the average concentration of Mn 5.94 mg/kg in the analyzed fish samples. The study revealed that the highest concentration of Mn was detected in Bigeye Ilisha at 21 mg/kg, while the lowest value was observed in Ribbon Fish at 1.1 mg/kg. In a study conducted by Mutlu et al. found that the concentration of Mn ranged between 0.35 mg/kg and 2.06mg/kg [56].

### Copper (Cu) in sampled sea fish (mg/kg)

The study found that the mean concentration of Cu in the fish samples was 7.29 mg/kg. The maximum concentration was found in Shrimp which was 11.5 mg/kg and the lowest value was found in Silver Pomfret which was 5.2 mg/kg. However, these values fall within the permissible limits established by the Joint FAO/WHO Expert report [57]. A Study in Malaysia found the concentration of Cu in Indian Mackerel 2.15 mg/kg which also persist below the permissible limit [58]. A study conducted in Tanzania, the concentration of Cu in Indian Mackerel ranged from 0.897 mg/kg to 3.953 mg/kg, with a calculated mean concentration of 2.734 mg/kg [59].

### Lead (Pb) in sampled sea fish (mg/kg)

The study reported a mean concentration of 9.70 mg/kg for Pb. The concentration of Pb displayed a range from 2.6 mg/kg in Fourfinger Threadfin to 14.4 mg/kg in Indian Mackerel, which exceeded the permissible limit established by the Food and Agriculture Organization (FAO) [47]. In a study conducted in Bangladesh, lower levels of Pb were identified in Loittya fish at 2.0 mg/kg and Ilish at 0.67 mg/kg compared to the lower level of Pb observed in this study in Fourfinger Threadfin at 2.60 and White Snapper at 3.90, both were below the MPL [60]. Another study conducted in Greece identified Pb concentrations in Longfin Tuna (0.557 mg/kg wet weight) that exceeded the permissible limits established by the European Union (0.3 mg/kg wet weight) [61].

The analysis exposed a positive correlation between Chromium and Manganese (0.587) which is statistically significant. This metal pair indicated the parameters were interconnected with each other and likely have a common origin within the study area [67, 68].

Strong positive correlations among heavy metals signify dual interdependence, emanation from similar sources, and same patterns in how they exert their influence [69, 70]. In a Bangladeshi study on Bombay Duck and Ilish found significant positive correlations between Cr and Pb (0.76) [60]. However, the majority of the heavy metals found in sea fish demonstrated non-significant correlations, some showed a negative correlation, which indicated different natural sources in sea fish samples.

The study assessed the day-to-day intake of specific heavy metals and subsequently compared these values with recommended guidelines to evaluate the safety of consuming sea fish samples, as presented in Table 3. Four EDI values for Cr exceeded the R_f_D of 0.0003 mg/kg, as indicated in the case of Ilish, Silver Pomfret, Longfin Tuna, and Bigeye Ilisha. Additionally, the study found that six estimated daily intake (EDI) values for Pb exceeded the R_f_D of 0.0004 mg/kg. These values were observed in Bombay Duck, Ilish, Silver Pomfret, Longfin Tuna, Indian Threadfin, and Bigeye Ilisha. The study reported the maximum recorded EDI value at 0.015139683 mg/kg-bw/day for Fe, which fell within an acceptable range relative to the reference dose. The lowest recorded EDI value was for Mn at 0.000316032 mg/kg-bw/day. A study shown in China on Bombay Duck and Silver Pomfret also identified a notably high EDI value for Chromium (Cr) [71].

**Table 3:** R_f_D and MPL of heavy metals for sea fish.

| Metal | R<sub>f</sub>D (mg/kg/d) | Reference | MPL (mg/kg/d) | Reference |
| --- | --- | --- | --- | --- |
| Fe | 0.667 | [36] | 100 | [52] |
| Ni | 0.020 | [62] | 80.000 | [25] |
| Mn | 0.140 | [33] | 20.000 | [63] |
| Cr | 0.003 | [64] | 0.600 – 4.350 | [47] |
| Pb | 0.004 | [65] | 2.170 | [47, 66] |
| Cu | 0.040 | [34, 62] | 30.000 | [57] |

**Table 4:** Pearson’s correlation among the heavy metals.

|  | Cr | Fe | Ni | Mn | Cu | Pb |
| --- | --- | --- | --- | --- | --- | --- |
| Cr | 1 |  |  |  |  |  |
| Fe | .111 | 1 |  |  |  |  |
| Ni | -.011 | .544 | 1 |  |  |  |
| Mn | **.587\*** | .106 | -.410 | 1 |  |  |
| Cu | -.098 | -.449 | .020 | -.188 | 1 |  |
| Pb | .524 | -.161 | .012 | -.021 | -.009 | 1 |

\* The correlation is statistically significant at the 0.05 level (2- tailed)

**Table 5:**
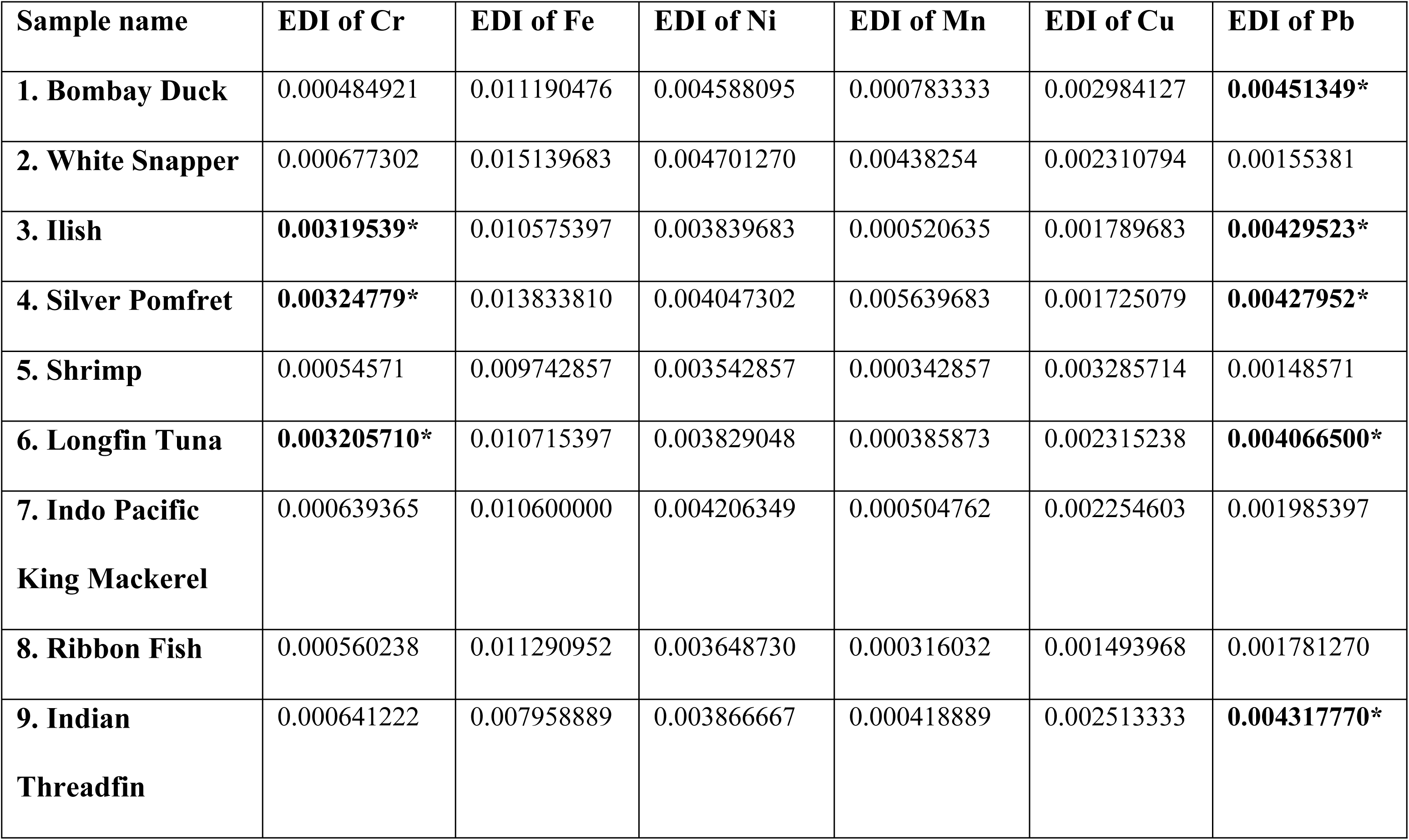
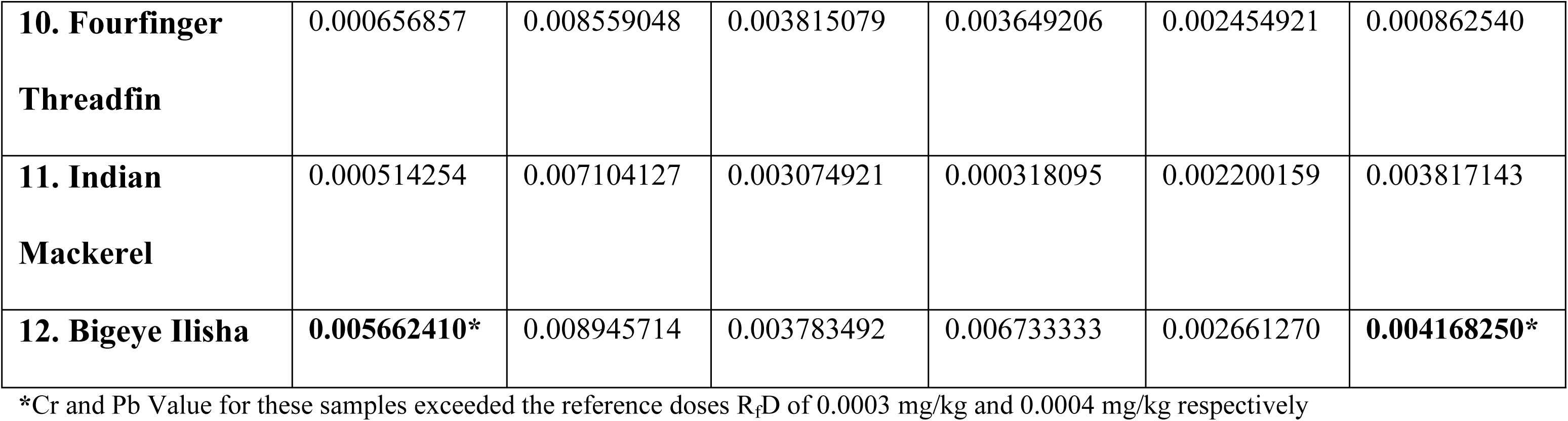
EDI (mg/kg-bw/day) of heavy metals through consumption of sampled sea fish.

| Sample name | EDI of Cr | EDI of Fe | EDI of Ni | EDI of Mn | EDI of Cu | EDI of Pb |
| --- | --- | --- | --- | --- | --- | --- |
| **1. Bombay Duck** | 0.000484921 | 0.011190476 | 0.004588095 | 0.000783333 | 0.002984127 | **0.00451349\*** |
| **2. White Snapper** | 0.000677302 | 0.015139683 | 0.004701270 | 0.00438254 | 0.002310794 | 0.00155381 |
| **3. Ilish** | **0.00319539\*** | 0.010575397 | 0.003839683 | 0.000520635 | 0.001789683 | **0.00429523\*** |
| **4. Silver Pomfret** | **0.00324779\*** | 0.013833810 | 0.004047302 | 0.005639683 | 0.001725079 | **0.00427952\*** |
| **5. Shrimp** | 0.00054571 | 0.009742857 | 0.003542857 | 0.000342857 | 0.003285714 | 0.00148571 |
| **6. Longfin Tuna** | **0.003205710\*** | 0.010715397 | 0.003829048 | 0.000385873 | 0.002315238 | **0.004066500\*** |
| **7. Indo Pacific King Mackerel** | 0.000639365 | 0.010600000 | 0.004206349 | 0.000504762 | 0.002254603 | 0.001985397 |
| **8. Ribbon Fish** | 0.000560238 | 0.011290952 | 0.003648730 | 0.000316032 | 0.001493968 | 0.001781270 |
| **9. Indian Threadfin** | 0.000641222 | 0.007958889 | 0.003866667 | 0.000418889 | 0.002513333 | **0.004317770\*** |
| **10. Fourfinger Threadfin** | 0.000656857 | 0.008559048 | 0.003815079 | 0.003649206 | 0.002454921 | 0.000862540 |
| **11. Indian Mackerel** | 0.000514254 | 0.007104127 | 0.003074921 | 0.000318095 | 0.002200159 | 0.003817143 |
| **12. Bigeye Ilisha** | **0.005662410\*** | 0.008945714 | 0.003783492 | 0.006733333 | 0.002661270 | **0.004168250\*** |

\*Cr and Pb Value for these samples exceeded the reference doses R<sub>FD</sub> of 0.0003 mg/kg and 0.0004 mg/kg respectively

### THQ and TTHQ

In table 8, the values of THQ and TTHQ of different heavy metals (Cr, Fe, Ni, Mn, Cu and Pd) are provided. These values are crucial for determining the adverse health outcomes associated with eating these fish because of the exposure to heavy metals. Within the table, it was detected that the THQ for Cr exceeded the established permissible limit of 1 in four fish samples (Ilish, Silver Pomfret, Longfin Tuna, Bigeye Ilisha). Furthermore, the THQ values for Pb surpassed the permissible limit of 1 in six fish samples (Bombay Duck, Ilish, Silver Pomfret, Longfin Tuna, Indian Threadfin, Bigeye Ilisha). The elevated value affirmed the existence of Cr and Pb contamination in the fish sample. Consistent ingestion of these polluted fish may end an elevated contact to Cr and Pb, resulting in various non-carcinogenic health risks for humans. For the remaining samples listed in the table, their THQ limits remained below the threshold of 1, indicating that the planes of these metals were inside acceptable values and did not pose a significant hazard to human health upon consumption.

Fish like Bombay Duck, Ilish, Silver Pomfret, Longfin Tuna, Indian Threadfin, and Bigeye Ilisha exceeded the THQ limit (>1) which indicated the residents may face potential non-carcinogenic health risks associated with the prolonged consumption of these fish. A recent study in Bangladesh found exceeded the THQ limit (>1) on Bombay Duck which was identified to pose a non-carcinogenic health risk [60]. Another Bangladeshi study estimated a higher THQ (1.646) value for Cr in the Ilish sample indicated potential health risk [72]. An study conducted by Praveena et al. also found the high THQ value (2.75) for Pb on Indian Mackerel which exceeded the THQ limit (>1) [58]. Whereas some studies investigated Shrimp, Longfin Tuna, Indian Threadfin, and Fourfinger Threadfin respectively and found the THQ value for Cr and Pb at safe levels (THQ < 1) [61, 73-75]

**Table 6:**
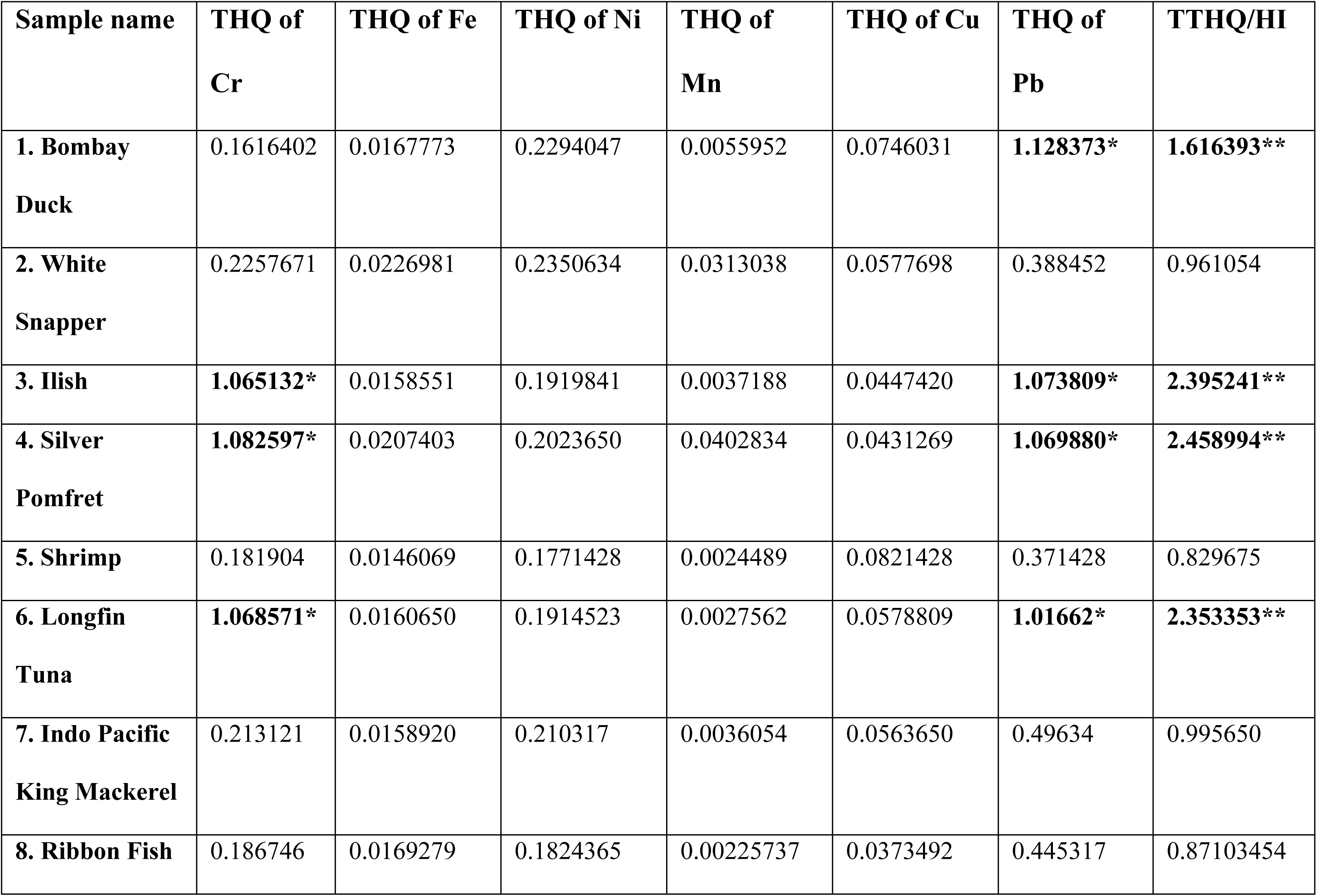
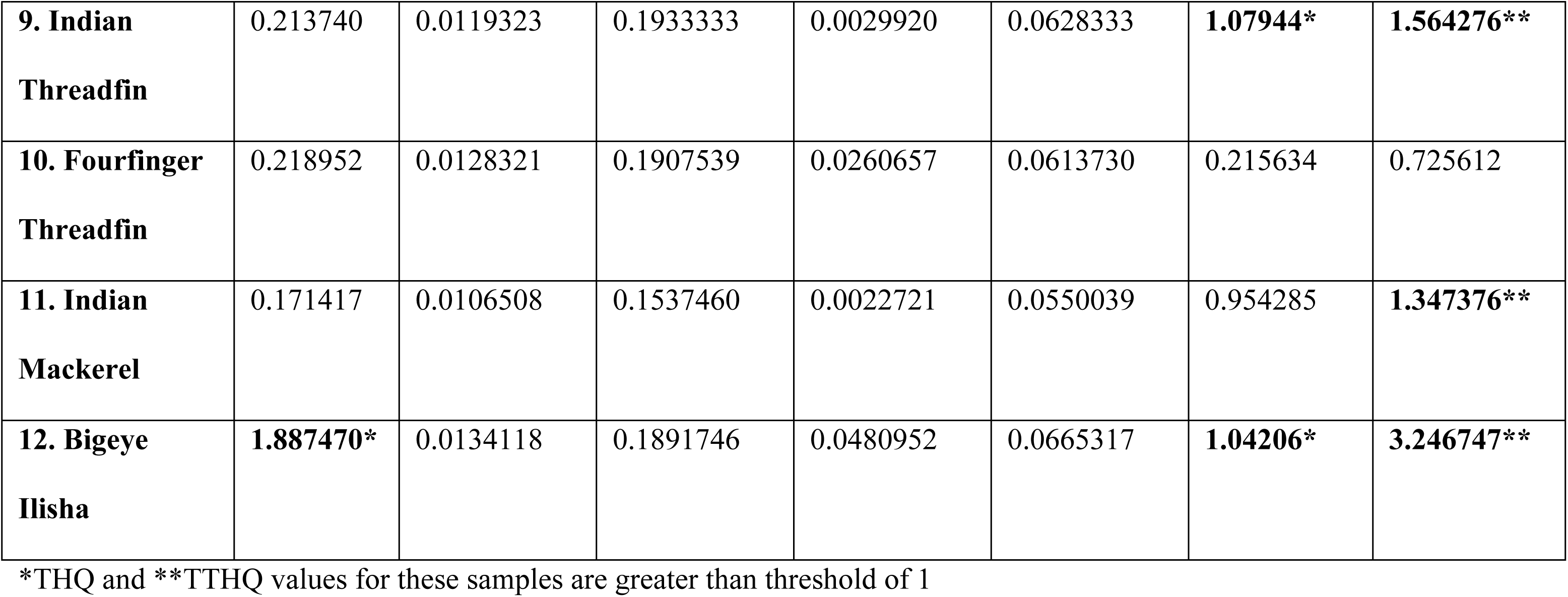
Target Hazard Quotient (THQ) and Total Target Hazard Quotient (TTHQ) of heavy metals through consumption of studied sea fish.

When the HI values surpasses a value of 1, it signifies that the respective metals are poisonous and pose a hazard to human well-being. However, in the current study, the HI values for five fish samples (White Snapper, Shrimp, Indo Pacific King Mackerel, Ribbon Fish, Fourfinger Threadfin) remained below the cut-off value of 1 suggested that the consumption of these fish did not pose a remarkable threat of metal-related hazards to the human body. But seven HI values in the sampled fish were greater than 1 clearly indicated a significant danger accompanying with the intake of these fish in terms of potential health hazards. The study identified the highest HI for heavy metals in Bigeye Ilisha (3.246747), while the lowest HI was observed in Indian Mackerel (1.3473766). A research found that if TTHQ/HI ratio for Longfin Tuna exceeded 1, it indicates an adverse health concern for those who consume this species [61]. A subsequent research studied by Alipour et al. indicated that Malaysian adults, as well as children in both Malaysia and Bangladesh, who intake Silver Pomfret over an extended period, face a significant non-carcinogenic health risk, as denoted by HI > 1 [76].

**Table 7:** Target Cancer Risk (CR) of heavy metals through ingestion of the sea fish samples.

| Sample name | CR of Cr | CR of Ni | CR of Pb |
| --- | --- | --- | --- |
| **1. Bombay Duck** | $1.98817 \times 10^{-5}$ | $7.79976 \times 10^{-6}$ | $3.83647 \times 10^{-5}$ |
| **2. White Snapper** | $2.77694 \times 10^{-5}$ | $7.99216 \times 10^{-6}$ | $1.32074 \times 10^{-5}$ |
| **3. Ilish** | **$1.31011 \times 10^{-4**}$** | $6.52746 \times 10^{-6}$ | $3.65095 \times 10^{-5}$ |
| **4. Silver Pomfret** | **$1.3316 \times 10^{-4**}$** | $6.88041 \times 10^{-6}$ | $3.6376 \times 10^{-5}$ |
| **5. Shrimp** | $2.23743 \times 10^{-5}$ | $6.02286 \times 10^{-6}$ | $1.26286 \times 10^{-5}$ |
| **6. Longfin Tuna** | **$1.31434 \times 10^{-4**}$** | $6.50938 \times 10^{-6}$ | $3.45653 \times 10^{-5}$ |
| **7. Indo Pacific King Mackerel** | $2.6214 \times 10^{-5}$ | $7.15079 \times 10^{-6}$ | $1.68759 \times 10^{-5}$ |
| **8. Ribbon Fish** | $2.29698 \times 10^{-5}$ | $6.20284 \times 10^{-6}$ | $1.51408 \times 10^{-5}$ |
| **9. Indian Threadfin** | $2.62901 \times 10^{-5}$ | $6.57333 \times 10^{-6}$ | $3.67011 \times 10^{-5}$ |
| **10. Fourfinger Threadfin** | $2.69311 \times 10^{-5}$ | $6.48563 \times 10^{-6}$ | $7.33159 \times 10^{-5}$ |
| **11. Indian Mackerel** | $2.10844 \times 10^{-5}$ | $5.22737 \times 10^{-6}$ | $3.24457 \times 10^{-5}$ |
| **12. Bigeye Ilisha** | **$2.32159 \times 10^{-4**}$** | $6.43194 \times 10^{-6}$ | $3.54302 \times 10^{-5}$ |

\*\* CR range lies in the Unacceptable range

The CR values identified in the selected fish samples ranged from 1.98817 × 10^-5^ to 2.32159× 10^-4^ for Cr; from 6.02286 × 10^-6^ to 7.99216 × 10^-6^ for Ni and from 1.26286 × 10^-5^ to 7.33159 × 10^-5^ for Pb. In general, a CR value exceeding 10^-4^ is considered unacceptable, while CR value ranging from 10^-4^ to 10^-6^ is deemed acceptable in terms of carcinogenic risk, and CR value below 10^-6^ is typically considered negligible, and value above 10^-1^ is considered very high probability of causing cancer [77]. In this study, the CR values for most of the heavy metals were in the acceptable range, and only four CR values for Cr lay in the unacceptable range. These were Ilish, Silver Pomfret, Longfin Tuna, and Bigeye Ilisha. Similarly, a Bangladeshi study conducted by Ali et al. showed that the CR value for Pb (2 ×10^-4^) on Bombay Duck was above the acceptable risk limit ( > 10^―4^) [60]. Another study in China conducted by Sarker et al., assessed the carcinogenic risk of sea fish for Pb (9.2 × 10^―7^) which was lower than the negligible level, whereas the value for Cr (2 × 10^―4^) was slightly exceeded the acceptable range for carcinogenic risk [71].

## Conclusions

The mean concentration of the selected heavy metals was observed in a descending order as follows: Fe (32.36) > Ni (12.12) > Pb (9.70) > Cu (7.29) > Mn (5.94) > Cr (5.22). Among the sea fish species, Chromium (Cr) and Lead (Pb) concentrations exceeded the acceptable levels established by international regulatory bodies such as the WHO and FAO. In this study, a high level of THQ was observed for Chromium (Cr) and Lead (Pb) among seven commonly consumed sea fish (Bombay Duck, Ilish, Silver Pomfret, Longfin Tuna, Indian Threadfin, Indian Mackerel, and Bigeye Ilisha) by people. The HI values also noted more than 1 among seven sea fish species reflecting the hazardous sign for human health. The CR of the sampled sea fish were in the unacceptable range for four fish samples (Ilish, Silver Pomfret, Longfin Tuna, and Bigeye Ilisha). Considering the health risks associated with elevated levels of heavy metals in sea fish, it is essential to ensure that the permissible limits of these metals in sea fish should not be exceeded. It is crucial to provide fishermen with education regarding the issues linked to excessive use of pesticides, insecticides, formalin, and other chemical additives or preservatives employed in sea fish. Public health awareness initiatives should be prioritized by the relevant regulatory bodies. A multifaceted strategy is required for these activities, involving the sharing of information through a variety of venues, including interactive workshops, social media campaigns, community seminars, and instructional scientific materials.

## Data Availability

The data necessary for the publication is already given in the manuscript, and if any further data is needed, it will be provided by the corresponding author.

